# Predicting the impact of primaquine on Plasmodium falciparum transmission using multiscale modelling

**DOI:** 10.64898/2026.10.21.26366166

**Authors:** Xiao Sun, Julie A. Simpson, James M. McCaw, Pengxing Cao

**Affiliations:** School of Mathematics and Statistics, The University of Melbourne, Melbourne, VIC, Australia; Department of Epidemiology and Biostatistics, Melbourne School of Public Health, The University of Melbourne, Melbourne, VIC, Australia; Nuffield Department of Medicine, University of Oxford, Oxford, UK

## Abstract

Primaquine is currently the only widely available antimalarial drug that targets mature *Plasmodium falciparum* gametocytes, the life stage in the human bloodstream that mediates transmission from humans to mosquitoes. The only study that provided large-scale field data suitable for examining the additional impact of the WHO-recommended dose of 0.25 mg/kg primaquine in an artemisinin-based combination therapy (ACT)-based mass drug administration (MDA) campaign was reported by Deng et al. (2018, Clinical Infectious Diseases). The study showed that both ACT alone and ACT plus primaquine substantially reduced average monthly malaria hospital rates and the prevalence of infection after the MDA campaign but no additional benefit was observed with primaquine. These findings challenge our expectations for the population-level impacts of primaquine and are in stark contrast to the predictions (of substantial population-level impacts) made by existing epidemiological-scale modelling studies. To address these knowledge gaps, we hypothesised that individuals’ adherence patterns to multi-round MDA campaigns could be a factor that explains these conflicting empirical observations. To examine this hypothesis, we developed a data-driven multiscale agent-based model of malaria infection and transmission, which quantitatively captures both the detailed biology of human infection kinetics and the pharmacodynamic actions of antimalarial drugs. Our model provides a coherent explanation for these apparently disparate observations, including the individual-level gametocytocidal effect of primaquine, the population-level impacts of primaquine in MDA campaigns, and the population-level impacts predicted by previous modelling studies. All three observations can be explained by underlying adherence patterns within a single modelling framework. We used our model to predict the effect of primaquine in routine case management outside MDA, finding that administration could significantly reduce the prevalence of infection, supporting its use as a transmission-blocking intervention against malaria.

## Introduction

Malaria is a vector-borne disease that threatens billions of people worldwide and causes approximately half a million deaths each year, with *Plasmodium falciparum* accounting for the greatest number of deaths among all human-infecting species [1]. Transmission of malaria parasites from humans to mosquitoes is mediated by the sexually committed life stage of the parasite within the human bloodstream, known as gametocytes. Their concentration within the blood-stream strongly affects the host’s infectiousness to mosquitoes [2]. Gametocytes are ingested by mosquitoes during blood feeding and subsequently develop into sporozoites that migrate to the salivary glands, enabling transmission back to humans, completing the cycle of infection and sustaining the parasite population within the environment. Efforts to suppress malaria transmission include a range of interventions targeting either the human host or mosquito vector, among which incorporation of gametocytocidal antimalarials into first-line therapies represents a promising strategy for interrupting the transmission cycle [3, 4].

Several antimalarial drugs target gametocytes, including primaquine, tafenoquine, and methylene blue [5–7], of which primaquine is the most widely available [8]. The World Health Organization (WHO) currently recommends a single low dose of 0.25 mg/kg primaquine in addition to standard artemisinin-based combination therapy (ACT) to reduce malaria transmission and support elimination efforts, particularly in low-transmission settings and regions approaching malaria elimination [1]. This recommendation is supported by clinical studies showing that a single low dose of primaquine substantially shortens the duration of gametocyte carriage in treated patients and rapidly reduces the probability of transmission from humans to mosquitoes [5, 6, 9–12]. The WHO-recommended dose also poses a low risk of serious adverse events, supporting its use without routine testing for glucose-6-phosphate dehydrogenase (G6PD) deficiency [8, 13].

Despite strong evidence of primaquine’s ability to block human-to-mosquito transmission, the population-level impact of primaquine remains under-studied. Several review articles have emphasised the need for large-scale cluster-randomised studies [14, 15]. However, conducting such studies is challenging for two main reasons. First, it is difficult to design studies with adequate control groups for cross-area comparisons to evaluate the population-level effect of primaquine. Specifically, key factors such as baseline transmission intensity, intervention coverage, and treatment adherence are difficult to control in field settings [16]. Second, in many endemic areas, multiple interventions are implemented concurrently, resulting in low or declining transmission intensity, making it difficult to detect any incremental effect of primaquine [17, 18]. To the best of our knowledge, the only study to date that provided large-scale field data suitable for examining the population-level transmission-blocking effect of the WHO-recommended 0.25 mg/kg dose of primaquine was conducted by Deng et al. [19]. In this study, two cohorts within the same area received different treatment regimens, irrespective of infection status, during a mass drug administration (MDA) campaign consisting of three monthly rounds. One cohort received ACT alone (artemisinin–piperaquine), while the other received ACT plus a 0.25 mg/kg dose of primaquine in each round of the MDA (with approximately 90% coverage achieved in both cohorts). Deng et al. found that the average monthly malaria hospital rates per 100,000 people (at 4–9 months post MDA) fell significantly and parasitemia rates in children also decreased substantially post MDA in both cohorts, but no additional benefit was observed with primaquine [19]. This finding highlights a knowledge gap between the well-established individual-level transmission-blocking effects of primaquine and the (to date) absence of a detectable impact at the population level.

Mathematical modelling plays a key role in elucidating the underlying reasons for empirical observations from epidemiological field studies. Two modelling studies have investigated the impact of primaquine on reducing the prevalence of *P. falciparum* in MDA campaigns. Maude et al. developed a transmission model to evaluate the contribution of adding primaquine to ACT-based intervention strategies, and predicted that the addition of primaquine would enhance the impact of both MDA and case management in reducing malaria prevalence, regardless of the presence or absence of artemisinin resistance [20]. Gerardin et al. used an agent-based transmission model (EMOD) to evaluate the effect of adding primaquine to ACT-based MDA strategies, predicting, just as Maude et al., that the addition of primaquine would further reduce malaria prevalence, particularly at higher coverage and when combined with long-acting prophylactic drugs [21]. These studies’ predictions are in stark contrast to the absence of a detectable impact observed in the field study by Deng et al. [19], suggesting that our understanding of the pharmacological and epidemiological impact of primaquine is incomplete.

One hypothesis for the discrepancy between model predictions and field observations is that both of these models [20, 21] failed to capture the adherence patterns observed in MDA settings [22]. Both models assumed that all individuals had an equal probability of receiving treatment during each round of MDA (i.e., independent participation across rounds); however, in practice, adherence across rounds is correlated; the probability of receiving treatment is strongly influenced by an individual’s willingness to participate in the programme. Adhikari et al. reported that approximately 90% of individuals participated in all three rounds of MDA campaigns [23], with the remaining 10% being partial or non-participants. Von Seidlein et al. also observed that approximately 57% of individuals chose to complete all three rounds of an MDA campaign [24]. Correlated behaviour between MDA rounds is well known (based on basic probability principles) to result in reduced population-level impact compared to uncorrelated behaviour [25, 26].

Beyond the likely contribution of correlated behaviour in participation in MDA programmes, additional details of the biology of infection and transmission—unaccounted for in existing modelling studies—are likely contributing to the apparent inconsistencies between both clinical and epidemiological empirical studies, and between modelling predictions and empirical field studies.

We address these apparent inconsistencies, through the development, calibration and evaluation of a detailed data-driven agent-based model of malaria infection that quantitatively captures the biological details of the parasite life cycle in both humans and mosquitoes. We ask whether individual’s adherence patterns to a multi-round MDA programme can explain the discrepancy between previous model predictions and field observations. We show that our model-based analysis provides a coherent explanation for the apparent discrepancies between the observed individual-level and epidemiological impacts of primaquine treatment. And we use our model to predict that the addition of primaquine, co-administered with standard ACT as part of a treatment strategy under routine case management, would substantially reduce the prevalence of infection, even when it has limited epidemiological impact when used in an MDA setting.

## Results

### The agent-based model of malaria transmission

We first provide an overview of the novel agent-based model, thereby establishing a clear context for understanding the analyses that follow, while leaving the full model details in the Materials and Methods. The agent-based model is a multiscale discrete-time stochastic model that integrates both population-level and within-host dynamics. At the population level, the model captures: (1) the state of each human individual, i.e., uninfected or infected (infected individuals can be symptomatic or asymptomatic) (Fig. 1, green block); (2) the number of mosquitoes in different classes, i.e., susceptible, exposed, or infectious (Fig. 1, grey block); and (3) the transmission between humans and mosquitoes. At the within-host level, the model captures (1) the dynamics of asexual parasites and gametocytes in human hosts (Fig. 1, blue block), coupled with pharmacokinetics–pharmacodynamics (PK–PD) models of antimalarial drugs to simulate time series of parasitemia and gametocytemia under different treatments (Fig. 1, purple block); and (2) a probability model describing gametocyte mating and production of zygotes within the mosquito midgut, underpinning the exposed state of mosquitoes (Fig. 1, light grey block).

**Figure 1:**
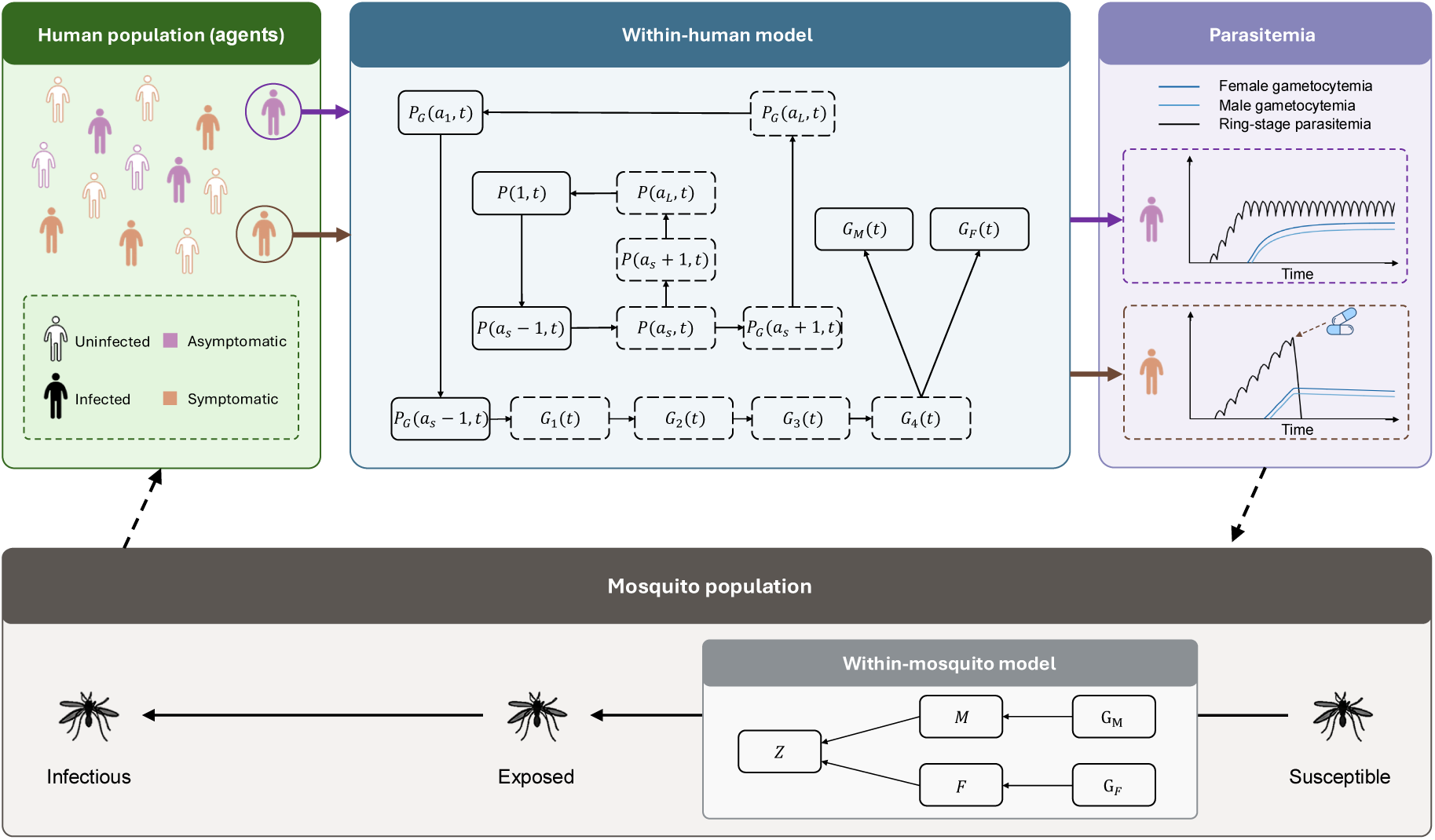
The multiscale agent-based model of malaria transmission. The agent-based population model consists of five coupled components: individual human agents, a within-host model, human-to-mosquito transmission, mosquito population dynamics, and mosquito-to-human transmission. For each individual in the human population, parasitemia and gametocytemia are generated by the within-host model. The mosquito population dynamics describe the progression of mosquitoes through susceptible, exposed, and infectious states. Human-to-mosquito and mosquito-to-human transmission link the human and mosquito populations by determining newly exposed mosquitoes and new human infections, respectively. Details of the model are provided in the Materials and Methods.

The multiscale agent-based model can simulate flexible adherence patterns for individuals participating in an MDA programme, overcoming the limitation of previous compartmental model structures, which implicitly assumed an equal probability of receiving treatment during each MDA round for each individual. Furthermore, the model extends upon previous agent-based models by explicitly incorporating both PK–PD effects of antimalarial drugs and a mechanistic, data-validated model of human-to-mosquito transmission [27]. This enables direct and explicit translation of our knowledge of the clinically observed individual-level effect of primaquine to the prediction of population-level impacts of primaquine-based intervention programmes.

### Single-dose primaquine markedly reduces the infectiousness of treated individuals

We first demonstrate that our model reproduces the clinical observation that adding the WHO-recommended 0.25 mg/kg dose of primaquine to a standard ACT (i.e., artemisinin–piperaquine), as used in the MDA study by Deng et al. [19], substantially shortens the duration of gametocytemia in treated individuals and reduces their infectiousness to mosquitoes [5, 6, 9–12]. This validation step is essential to ensure that our agent-based model is grounded in accurate individual-level behaviour, and to rule out the possibility that any non-significant drug efficacy observed at the population level in our model is due to a poor representation of individual-level efficacy. The within-human model (Fig. 1, blue block) was used to simulate antimalarial PK profiles and parasite kinetics within an individual. We used published PK–PD models of dihydroartemisinin (DHA), an artemisinin derivative, and piperaquine (PPQ) to simulate the standard ACT [28, 29]. The individual’s infectiousness quantifies the probability that a susceptible mosquito becomes exposed following a blood meal.

The simulated time series of plasma concentrations of antimalarial drugs (DHA, PPQ, and primaquine), together with the corresponding parasitemia, gametocytemia, and the individual’s infectiousness to mosquitoes, are shown in Fig. 2. The ACT, comprising 120 mg DHA and 960 mg PPQ given at 0, 24, and 48 hours post-treatment initiation (i.e., days 15, 16, and 17 post-infection) according to WHO guidelines [1], was administered on day 15 post blood-stage infection (Fig. 2a), when gametocytemia is approximately 10^4^ per mL, consistent with levels observed in previous clinical trials [5, 6]. For the scenario where primaquine was administered in addition to ACT, a single dose of 0.25 mg/kg primaquine was given at the same time as the first dose of ACT (the representative PK profile was simulated for an individual with a body weight of 60 kg). Fig. 2b shows the corresponding simulated time series for parasitemia and gametocytemia under ACT alone and ACT combined with 0.25 mg/kg primaquine. Compared with ACT alone, the addition of a single dose of 0.25 mg/kg primaquine leads to a rapid reduction in gametocytemia of approximately one order of magnitude. This substantial reduction in gametocytemia translates into a corresponding model-predicted reduction in the individual’s infectiousness to mosquitoes (Fig. 2c) of up to approximately 90%. Our model predictions are consistent with clinical data reported in [5]. In the ACT alone group, the infectiousness of infected individuals on day 2 post-treatment was similar to, or higher than, that at treatment initiation, whereas the addition of 0.25 mg/kg primaquine led to an approximate 93% reduction in median infectiousness on day 2 post-treatment. These results demonstrate that our within-human model, coupled with PK–PD models of antimalarial drugs (DHA, PPQ and PQ), captures the effect of primaquine in reducing individual infectiousness, providing a solid foundation for using our agent-based model to evaluate the population-level impact of primaquine on malaria transmission.

**Figure 2:**
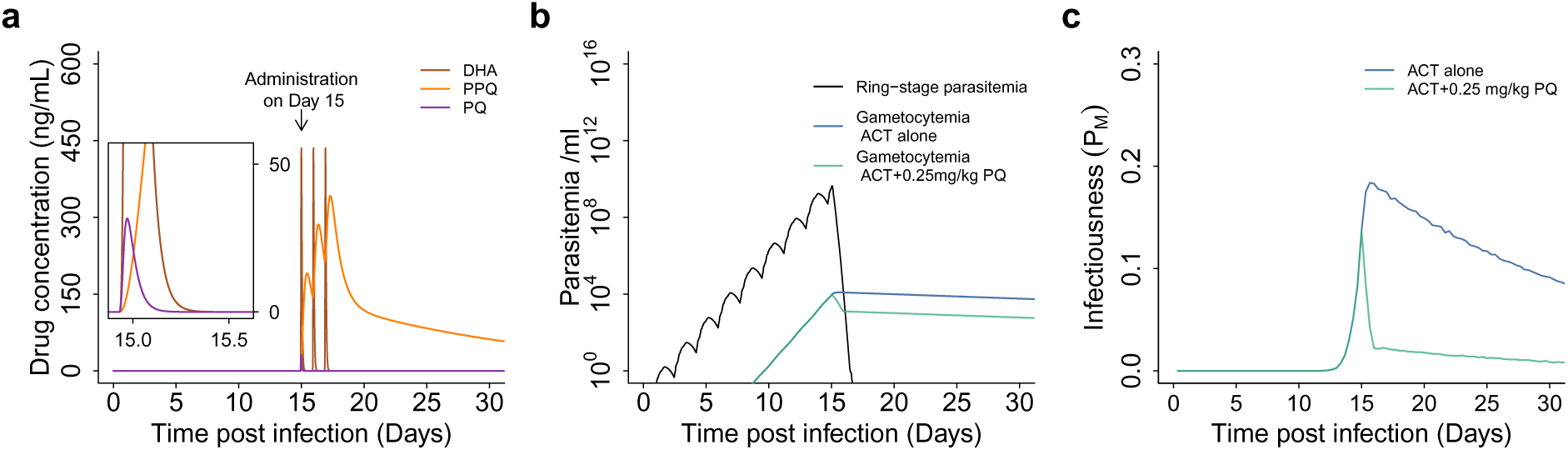
Effect of the addition of 0.25 mg/kg primaquine to ACT on parasite dynamics and infectiousness. *P_M_*. (a) Pharmacokinetic (PK) profiles of dihydroartemisinin (DHA, brown curve), piperaquine (PPQ, orange curve) and primaquine (PQ, purple curve) with treatment administered on Day 15 post blood stage infection. The inset shows a zoomed-in view of PQ concentration. (b) Simulated trajectories of parasitemia (black curve) and gametocytemia under ACT alone (blue curve) and ACT combined with 0.25 mg/kg primaquine (green curve). (c) The infectiousness *P_M_*under ACT alone (blue curve) and ACT combined with 0.25 mg/kg primaquine (green curve).

### Population-level impact of primaquine in MDA campaigns depends on adherence

We simulate an MDA campaign applied to a hypothetical human population of 1000 individuals using our agent-based model. The MDA campaign comprises three rounds at monthly intervals, using either ACT alone or ACT combined with a single 0.25 mg/kg dose of primaquine, with 90% population coverage in each round, consistent with the design of Deng et al. [19]. We simulate the MDA campaign under three adherence scenarios: (1) full adherence, in which a fixed 90% of individuals are selected to participate in all three rounds; (2) half adherence, in which a fixed 45% of individuals are selected to participate in all three rounds and the other 45% are independently selected in each round to maintain 90% coverage; and (3) independent participation across rounds, in which 90% of individuals are independently selected to participate in each round. Previous modelling studies [20, 21] have assumed independence (i.e., scenario three). For each scenario, we simulate 100 pairs of stochastic MDA simulations, with each pair consisting of one simulation in which no PQ is administered (i.e., ACT alone) and the other in which a single dose of 0.25 mg/kg PQ is administered (i.e., ACT+PQ 0.25 mg/kg).

The simulated prevalence of infection—defined as the proportion of the population with blood-stage parasites—under ACT alone and ACT combined with 0.25 mg/kg primaquine under the three adherence scenarios is shown in Fig. 3. Based on the median curves of the model-predicted prevalence (ACT alone: blue curves; ACT+PQ: green curves), we find that our model is able to capture both the field observations reported by Deng et al. [19], and the predictions made by previous modelling studies [20, 21]. Specifically, under full adherence (Fig. 3a), the addition of 0.25 mg/kg primaquine to a standard ACT (i.e., DHA+PPQ) produces no material additional reduction in prevalence compared to ACT alone. However, as the adherence level decreases, the additional reduction in prevalence (over a period of up to 18 months post MDA) induced by the addition of primaquine becomes increasingly observable (Fig. 3b and c). A similar trend is observed in the modelled population-averaged gametocytemia and infectiousness to mosquitoes (Supplementary Figure 1), underpinning the changes in the observed prevalence of infection. A paired analysis, in which the difference between each pair of simulated prevalence curves was calculated (Fig. 3d–f), quantifies the variation in the absolute reduction in prevalence, and indicates that, under ideal circumstances (Fig. 3c), maximum impact would be expected around 9–12 months post MDA.

**Figure 3:**
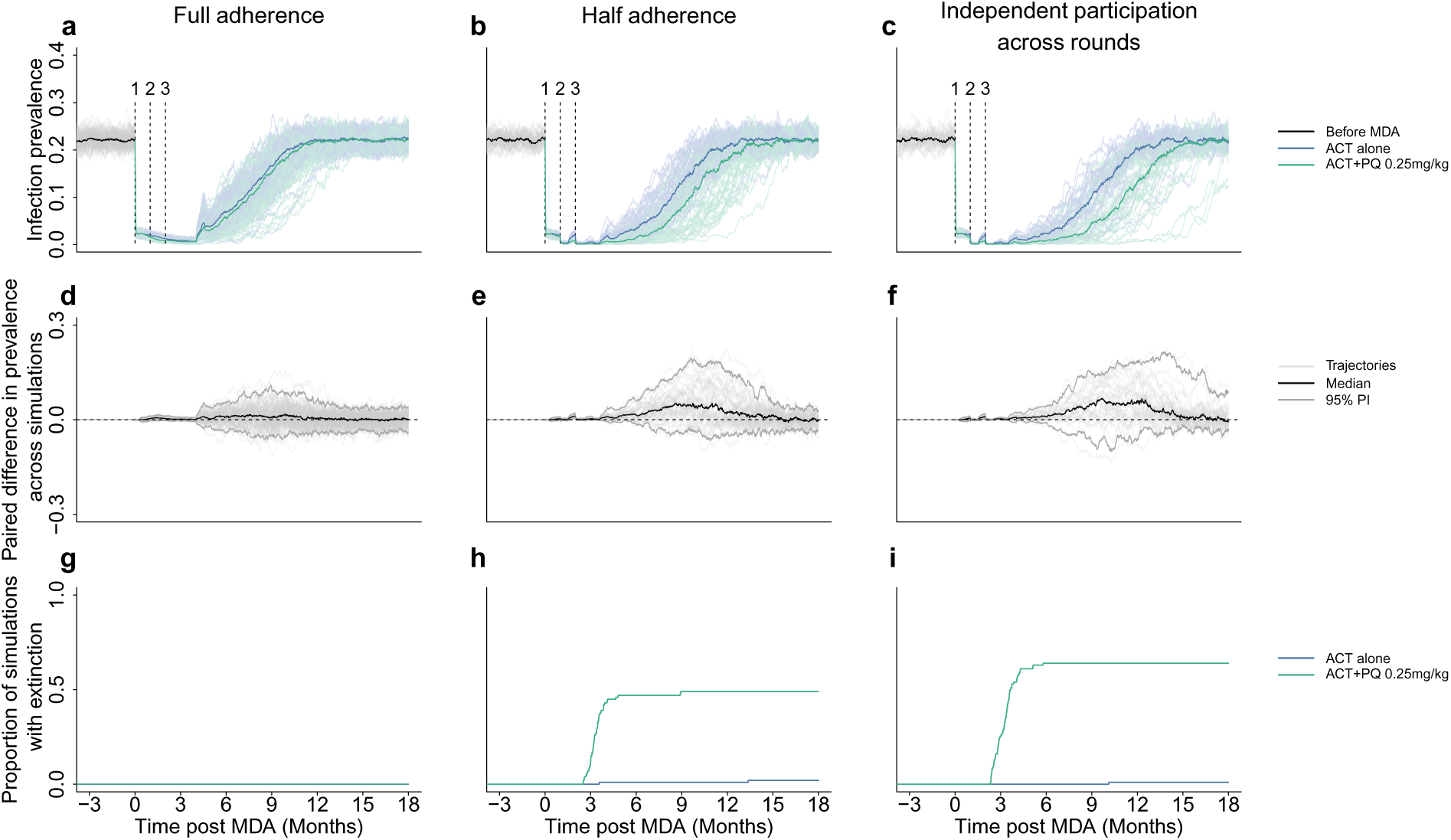
Effect of adding a single dose of 0.25 mg/kg primaquine to ACT on the prevalence of infection during MDA under three adherence scenarios. Three scenarios are considered: full adherence (left panels, a, d and g) in which the same 90% of participants receive treatment across all three rounds and 10% are never treated, half adherence (middle panels, b, e and h) in which which 45% of participants receive treatment across all three rounds and an additional 45% are selected at random for each round, and independent participation across rounds (right panels, c, f and k) in which there is no correlation across rounds in terms of which 90% of individuals receive treatment. For each scenario, we simulated 100 pairs of stochastic MDA simulations, with each pair consisting of one simulation in which no PQ was administered (i.e., ACT alone, blue) and another in which a low dose of 0.25 mg/kg PQ was administered (i.e., ACT+PQ 0.25 mg/kg, green). Panel (a) shows model-simulated stochastic trajectories of the prevalence of infection for the scenario with full adherence. The black curve represents the median prevalence prior to MDA (individual simulated prevalence trajectories are shown in grey), while the coloured curves represent the median prevalence during and after MDA (with individual simulated prevalence trajectories shown by lighter curves). Blue represents treatment without PQ (i.e., ACT alone), and green represents ACT combined with 0.25 mg/kg primaquine (ACT+PQ 0.25 mg/kg). The numbered vertical dashed lines indicate the timing of the three monthly MDA rounds. Panel (b) shows 100 model-simulated stochastic trajectories of the prevalence of infection for the scenario with half adherence. Panel (c) shows 100 model-simulated stochastic trajectories of the prevalence of infection for the scenario with independent participation across rounds. Panels (d)–(f) show the difference in prevalence between paired simulations for the corresponding scenarios from (a)–(c). The black curve represents the median difference in prevalence, and the upper and lower grey curves represent the 95% prediction intervals (PIs) of the paired prevalence differences, calculated from the individual differences within each pair (shown as thin, light grey curves). All trajectories and the median shown in (a)–(f) are conditional on non-extinction. Panels (g)–(i) show the cumulative proportion of simulations resulting in stochastic extinction of both infected humans and infected mosquitoes under the three scenarios.

While it follows from fundamental probabilistic principles that increasing individual-level adherence in an MDA programme, while keeping overall coverage fixed, will result in reduced coverage of infected individuals and, consequently, a reduced impact on the prevalence of infection, this relationship alone does not imply that the predicted impact of the addition of primaquine under a full adherence scenario is non-significant. Adherence information was not reported in the study by Deng et al. [19]. Based on other studies that have reported adherence was generally above 50% and as high as 90% [23, 24], our results suggest that adherence in the trial reported by Deng et al. was likely to be much higher than 50% and closer to 100%.

An MDA programme may also drive local extinction, where the number of both infected humans and mosquitoes is zero. Extinction does not occur (Fig. 3g) or is extremely rare (Fig. 3h,i) under standard ACT. The addition of primaquine dramatically increases the proportion of simulations resulting in extinction under scenarios with half and independent participation across rounds (Fig. 3h,i). In contrast, no extinction events were observed under the full-adherence scenario, in which 10% of the population never receive treatment and hence maintain a reservoir for re-establishment of transmission in the population. These anticipated effects of primaquine when delivered as part of MDA, and the influence of adherence (i.e., the correlation structure of who receives treatment across rounds) would be infeasible to assess in field trials; it would require a large number of replications of the entire MDA programme under nearly identical conditions. Our results strongly suggest that, in small populations (e.g., on the order of 1000 individuals) and with independent participation across rounds, adding primaquine to an ACT-based MDA would be able to drive local extinction in around 50% of deployments, in contrast to ACT-only MDA in which extinction is highly unlikely.

### Sustained transmission reduction when primaquine is included in routine case management

We now consider a scenario in which 0.25 mg/kg primaquine is co-administered with standard ACT as part of a treatment strategy under routine case management. Unlike MDA campaigns, which are implemented over a limited period, routine case management is provided on an ongoing basis and therefore represents a sustained intervention targeting malaria transmission in endemic settings [30]. We illustrate the potential impact of primaquine by comparing two scenarios: (1) ACT alone, where ACT (i.e., DHA+PPQ) is used alone as the routine treatment, and (2) ACT+PQ, where a single dose of 0.25 mg/kg primaquine is introduced alongside ACT. Details of the simulations are provided in the Materials and Methods. Following the introduction of 0.25 mg/kg primaquine, the prevalence of infection decreases rapidly (compared to prevalence under ACT alone) and approaches a new epidemiological equilibrium (Fig. 4a). This reduction in the prevalence of infection is driven by the individual-level impact of primaquine on human-to-mosquito infectiousness (recall Fig. 2c) and is evident in the clear declines in population-averaged gametocytemia (Fig. 4b) and infectiousness to mosquitoes (Fig. 4c) under ACT+PQ.

**Figure 4:**
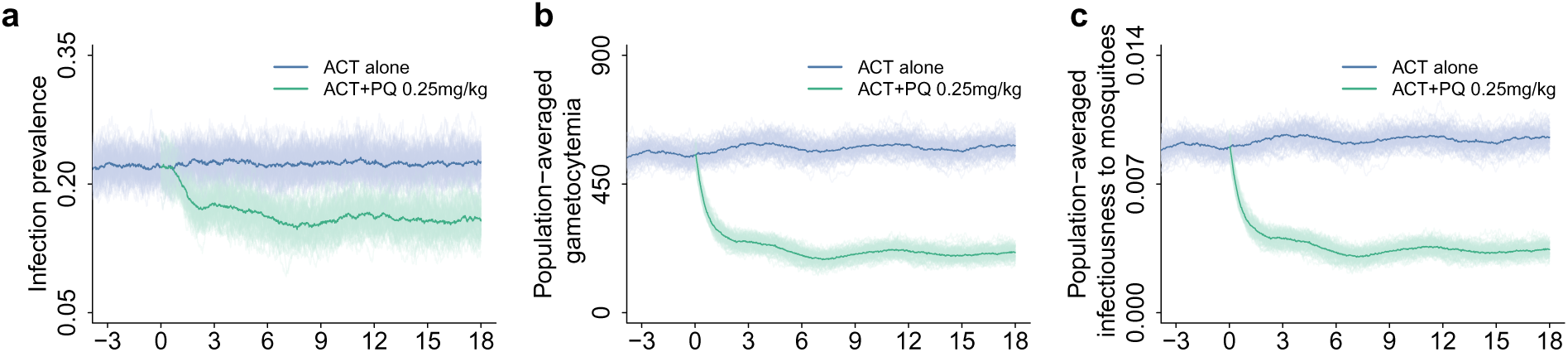
Effect of introducing a single dose of 0.25 mg/kg primaquine to ACT under routine case management. Model simulated stochastic trajectories of the prevalence of infection under routine case management with and without primaquine are shown in (a). Coloured curves represent the median prevalence of infection (with individual simulated prevalence trajectories shown by lighter curves). Blue represents treatment without PQ (i.e., ACT alone), and green represents ACT combined with 0.25 mg/kg primaquine (ACT+PQ 0.25 mg/kg). Primaquine treatment is introduced from a specified time (labeled 0). The population size was 1000, and 100 stochastic simulations were generated for each scenario (i.e., ACT alone versus ACT+PQ 0.25 mg/kg). Corresponding trajectories of population-averaged gametocytemia (i.e., the mean gametocytemia across the 1000 individuals) are shown in (b), and of population-averaged infectiousness to mosquitoes (i.e., the mean probability of transmission from the 1000 individuals to mosquitoes) in (c).

## Discussion

In this study, we developed a novel agent-based model of malaria transmission that integrates PK–PD models of antimalarials and a mechanistic, data-validated human-to-mosquito transmission model within a population-scale framework to evaluate the impact of primaquine on blocking malaria transmission and reducing prevalence. By simulating the model at both the individual and population levels, we showed that the model reproduced not only the significant gametocytocidal effect of primaquine observed in clinical studies, but also its non-significant impact on reducing malaria prevalence in the only MDA study by Deng et al. [19] designed to evaluate the benefit of a 0.25 mg/kg dose of primaquine co-administered with ACT. This provides a plausible explanation for these seemingly conflicting observations. Through simulations of scenarios with distinct assumptions about participant adherence in MDA, we further provide an explanation for why predictions from previous models that assume, either explicitly or implicitly, independent participation across rounds, have consistently suggested a significant benefit of including primaquine in MDA [20, 21], whereas field studies, such as that of Deng et al. [19] suggest no additional benefit from the inclusion of primaquine.

Our agent-based model allows us to explore the underlying processes that should, based on biological first principles and clinical efficacy studies, influence how drug administration affects the prevalence of infection. For example, under MDA, the effectiveness of primaquine in reducing the prevalence of infection is predicted to depend on the effective coverage of drug administration (Figs. 3a–c): the smaller the proportion of individuals who never receive primaquine treatment, the greater its effect on reducing the prevalence of infection. Specifically, for the simulated three-round MDA with 90% coverage in each round, the effective (one-dose) coverage is 90% under the full adherence scenario, compared with 99.9% ((1 *−* (1 *−* 0.9)^3^) *×* 100%) under independent participation across rounds, with corresponding notable differences in modelled impact. Furthermore, we found that the predicted impact of primaquine was strongly associated with reductions in population-averaged gametocytemia and, in turn, with population-averaged infectiousness to mosquitoes immediately following administration of the three MDA rounds (Supplementary Figure 1). These results, combined with those from our analysis of adding primaquine to routine case management (Fig. 4), confirm the fundamental role that gametocytemia concentration plays in determining the infectiousness of humans to mosquitoes (Fig. 2, and [27, 31]) and so shaping the prevalence of infection.

We simulated three specific scenarios of participation across MDA rounds. Full adherence assumes that individuals who participate in the first round continue to participate in all subsequent rounds, whereas those excluded remain non-participants throughout. Under half adherence, half of the individuals covered in each round participate consistently across all rounds, while the remaining coverage is filled by individuals independently selected in each round. Independent participation across rounds assumes no correlation in participation between rounds. Although these three scenarios are sufficient to demonstrate the importance of adherence in determining the impact of primaquine, in practice, participation behaviour is difficult to control. This is because it is shaped by a combination of persistent and transient factors: some individuals may be consistently excluded, for example, because of illness or pregnancy, whereas others may miss rounds intermittently because of mobility, such as travel or participation fatigue over repeated rounds [32, 33]. Correlated participation behaviour across treatment rounds in MDA has been modelled for other infectious diseases [25, 26] and could be readily incorporated into our agent-based model to simulate more complex malaria MDA scenarios, provided that individual-level adherence behaviours are specified.

In addition to the individuals’ participation behaviour in MDA, our model could be extended to study the dynamics of malaria transmission in other scenarios. For example, partial immunity, which develops over time through repeated infections, can suppress parasite growth and prevent parasite densities from reaching levels sufficient to cause clinical symptoms [34, 35]. To study how the pattern of reinfections may influence the long-term prevalence of malaria infection, we could extend our model by allowing the proportion of asymptomatic infections to vary depending on the number of repeated exposures to *P. falciparum* (as opposed to a fixed parameter *θ* assumed in the current study). Furthermore, our model could be extended to incorporate different intervention strategies and economic constraints, thereby enabling evaluation of the cost-effectiveness of alternative intervention strategies [36–38].

Our analyses indicate that adding 0.25 mg/kg primaquine to routine case management could provide a substantial transmission-blocking benefit (Fig. 4), supporting its potential as a valuable intervention for malaria control. Our predictions are consistent with a previous simulation study by Eziefula et al. [39], despite differences in model structure and parameterisation. Specifically, the model used by Eziefula et al. incorporated the effects of primaquine into an agent-based model [40, 41] by *assuming* that primaquine reduces the duration of infection by 78% and the level of infectiousness by 67% for each (and every) individual treated [39]. In contrast, our model incorporates a detailed data-driven PK–PD model of primaquine to capture its effects on gametocyte dynamics within individuals. Primaquine’s effects on transmission are therefore an emergent property of our multiscale model, rather than an assumed effect incorporated into the model. Eziefula et al. reported a substantially greater benefit of introducing primaquine in low-transmission settings than in high transmission settings [39]. Our parameterisation includes several features relevant to low-transmission settings, including within-human parameters derived from a cohort in such a setting [27, 29] and a modelled relatively low proportion of asymptomatic infections (i.e., 0.05), which is typically observed in low-transmission settings [42, 43]. These features support the qualitative consistency between our model predictions and those of Eziefula et al.

In conclusion, we have developed a multiscale modelling framework that has allowed us to reconcile the seemingly conflicting observations at the individual and population levels, as well as discrepancies between previous model predictions and field observations. This represents a major advance in our understanding of malaria transmission dynamics and the effects of antimalarial interventions. Our study provides a quantitative, mechanistic evaluation of the transmission-blocking effects of primaquine under both MDA and routine case management, and establishes a framework for predicting the clinical and public health impacts of transmission-blocking interventions.

## Materials and Methods

### Agent-based model of malaria transmission

The agent-based model is a multiscale discrete-time stochastic model that integrates both population-level and within-host dynamics (Fig. 1). At the population level, the model captures: (1) the state of each human individual, i.e., uninfected or infected (infected individuals can be symptomatic or asymptomatic) (Fig. 1, green block); (2) the number of mosquitoes in different classes, i.e., susceptible, exposed, or infectious (Fig. 1, grey block); and (3) the transmission between humans and mosquitoes. At the within-host level, the model captures (1) the dynamics of asexual parasites and gametocytes in human hosts (Fig. 1, blue block), coupled with PK–PD models of antimalarial drugs to simulate time series of parasitemia and gametocytemia under different treatments (Fig. 1, purple block); and (2) a probability model describing gametocyte mating and production of zygotes within the mosquito midgut, underpinning the exposed state of mosquitoes (Fig. 1, light grey block). Details of the different model components and their mathematical formulations are provided in the following sections.

### Population level

A hypothetical human population is modelled by *N_H_* individual agents. Agents are classified by infection status as uninfected, asymptomatically infected, or symptomatically infected, as shown in Fig. 1. Following successful infection after a mosquito bite, an uninfected agent becomes infected. Whether the individual develops symptoms is a result of many factors such as immune responses or infection history, and we use a binary parameter, *ψ*, to indicate whether the infection of the individual is symptomatic (*ψ* = 0) or asymptomatic (*ψ* = 1). Given a proportion *θ* of *N_H_*agents developing asymptomatic infections, the values of *ψ* for all agents are deterministically specified to be *ψ* = 1 for *θN_H_* agents and *ψ* = 0 for the remaining (1*−θ*)*N_H_* agents.

For each infected agent, a within-human model is used to simulate within-host parasite dynamics, including blood-stage parasite maturation and replication, as well as the development of female and male gametocytes in the symptomatic and asymptomatic individuals. Asymptomatic and symptomatic infections can exhibit different profiles of parasitemia (the number of asexual parasites per mL of blood), thereby generating different profiles of female and male gametocytemia (the number of female and male gametocytes per mL of blood), which in turn influence the probability of transmission from infected agents to mosquitoes. Details of the within-human model are introduced in the following section.

The mosquito population of size *N_V_* is comprised of three compartments: susceptible (*S_V_*), exposed (*E_V_*), and infectious (*I_V_*) mosquitoes, as in the traditional Ross–Macdonald model [44]. Susceptible mosquitoes may become exposed after taking blood meals from infectious humans, and exposed mosquitoes subsequently become infectious after completing the extrinsic incubation period. At each time step (Δ*t* = 1 hour), the mosquito population is updated to account for mosquito births, natural deaths, new infections, and progression from the exposed to the infectious state, as follows:

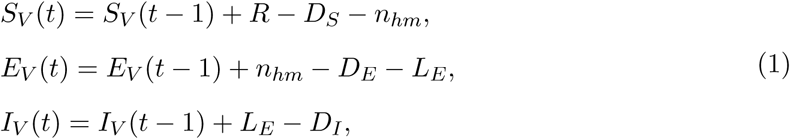

where *R* represents the number of mosquito births, sampled from a Poisson distribution Poi(Λ*_V_* Δ*t*), and Λ*_V_* is the mosquito birth rate. Here, we set Λ*_V_* = *N_V_ µ_v_*, where *µ_v_* is the mosquito death rate and *N_V_* is the mosquito population size, such that the mosquito population fluctuates over time with mean *N_V_*. The numbers of susceptible mosquitoes, *D_S_*, exposed mosquitoes, *D_E_*, and infectious mosquitoes, *D_I_*, that die during each time step are sampled from Poi(*µ_v_S_V_* (*t −* 1)Δ*t*), Poi(*µ_v_E_V_* (*t−*1)Δ*t*), and Poi(*µ_v_I_V_* (*t−*1)Δ*t*), respectively. The number of susceptible mosquitoes becoming exposed, *n_hm_*, is determined by the number of susceptible mosquitoes that take a blood meal from humans and whether at least one zygote develops in the midgut during each blood meal. To determine *n_hm_* in each time interval, we first calculate the number of bites on humans by susceptible mosquitoes *n_S_*, as follows:

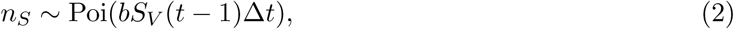

where *b* is the mosquito biting rate. In our model, the mosquito biting rate is assumed to be the inverse of the duration of the gonotrophic cycle, defined as the time required for host-seeking, blood-feeding, digestion, egg maturation, and egg-laying [45]. Although mosquitoes may continue host-seeking and take multiple bites within a single gonotrophic cycle until sufficient blood is acquired, each gonotrophic cycle is assumed in the model to correspond to the acquisition of a complete blood meal from a single human host.

For each susceptible mosquito taking a blood meal from a human, indexed by *i* = 1*, …, n_S_*, the bitten individual *h_i_*is selected at random from the human population:

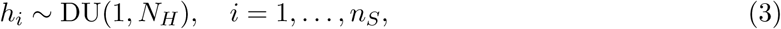

where DU(1*, N_H_*) represents the discrete uniform distribution on *{*1*, …, N_H_}*. Whether mosquito *i* becomes exposed during the blood meal is then determined by a human-to-mosquito transmission model that we previously developed [27]. The model captures both the transfer of female and male gametocytes from human blood to the mosquito midgut during the blood meal and the subsequent development of zygotes within the mosquito midgut.

Denoting the male and female gametocytemia of the bitten individual *h_i_* by *G_M,hi_* and *G_F,hi_*, respectively (the calculations of male and female gametocytemia are described in the next section, which focuses on the within-human level), following [27], the number of male gametocytes ingested by mosquito *i* during the blood meal from individual *h_i_*, denoted by *M_i_*, follows:

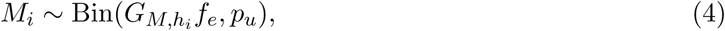

where *f_e_* is the fraction of the total human blood volume *V_H_* localised to the site of a mosquito bite, with *V_H_* assumed to be 4000 mL. Assuming that circulating male gametocytes are homogeneously distributed throughout the blood, the maximum number of male gametocytes available for uptake in a blood meal is given by *G_M,h__i_f_e_*. The probability of gametocyte uptake, *p_u_*, is given by *p_u_* = *V/*(*f_e_V_H_*), the ratio of the mosquito blood meal volume *V* to the volume of blood *f_e_V_H_*that contains *G_M,h__i_f_e_* male gametocytes. The number of female gametocytes ingested by mosquito *i* during the blood meal *F_i_* is defined similarly:

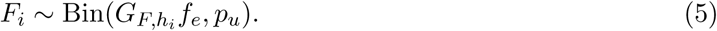

Each female gametocyte gives rise to a single female gamete, whereas each male gametocyte may produce *ρ* viable male gametes through replication (see [27] for details). The number of zygotes formed in mosquito *i*, denoted by *Z_i_*, is modelled as

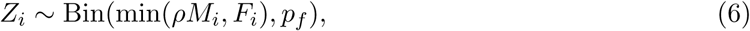

where *ρ* is the male gamete-to-gametocyte ratio and *p_f_*is the probability of successful fertilization for each pair of viable gametes. The parameter *ρ* represents an aggregate quantity combining the fraction of male gametocytes that produce male gametes through replication and the number of viable male gametes produced per replicating male gametocyte. A mosquito *i* is considered infected if at least one zygote forms. The infection status is therefore defined by the indicator function:

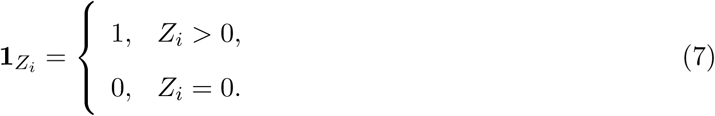

The total number of susceptible mosquitoes becoming exposed during the time step is then obtained by summing the infection indicators over the *n_S_* susceptible mosquitoes that take a blood meal:

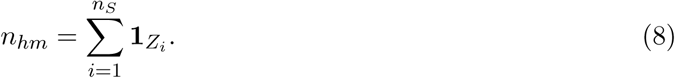

The number of exposed mosquitoes that progress to the infectious state, *L_E_*, is sampled from Poi(*E_V_* (*t −* 1)*k_v_*Δ*t*), where *k_v_*is the progression rate from the exposed state (i.e., developing zygotes) to the infectious state (i.e., developing sporozoites). Note that the model does not explicitly capture the timing or number of sporozoites developing within the mosquito. Instead, it uses *k_v_* to quantify the timing of mosquitoes becoming infectious, implicitly accounting for sporozoite development.

We next model transmission from infectious mosquitoes to humans. The number of bites on humans by infectious mosquitoes during each time step *n_I_*is given by:

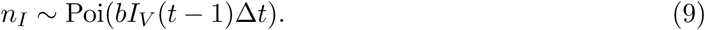

The number of successful mosquito-to-human transmission events *n_mh_* is then given by:

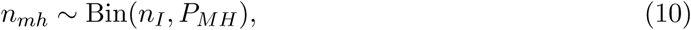

where *P_MH_* is the probability of mosquito-to-human transmission per infectious bite. For each successful mosquito-to-human transmission event, indexed by *j* = 1*, …, n_mh_*, the infected individual *h_j_* is selected at random from the human population:

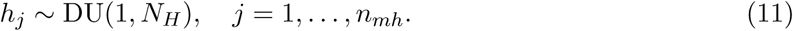

For individual *h_j_*, after a liver-stage infection period, *l_h_*, asexual blood-stage parasites are established and initiate successive asexual replication cycles. The initial number of asexual blood-stage parasites *P*_0_(*a*), which depends on parasite age *a*, is given by:

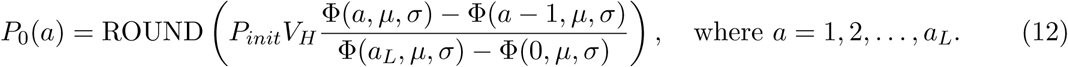

*P_init_* is the initial asexual parasite concentration. *V_H_* is the human blood volume. Φ(*a, µ, σ*) denotes the cumulative distribution function (CDF) of a normal distribution with mean *µ* and standard deviation *σ*, evaluated at *a*. *a_L_* is the duration (in hours) of each asexual replication cycle. The function ROUND(*·*) rounds the calculated value to the nearest integer.

### Within-human level

At the within-human level, we developed a PK–PD model by integrating published PK–PD models for dihydroartemisinin (DHA) [28], piperaquine (PPQ) [29], and primaquine (PQ) [46] with our previously developed within-host parasite dynamics model [27]. PPQ was selected as the ACT partner drug to ensure consistency with the treatment regimen used in the field study by Deng et al. [19]. The resulting model was used to capture drug-mediated parasite dynamics in infected individuals.

The model contains three major parasite populations: asexual parasites, *P* (*a, t*); sexually committed parasites, *P_G_*(*a, t*); and gametocytes, comprising immature stages *G*_1_(*t*)–*G*_4_(*t*) and mature male and female gametocytes, *G_M_* (*t*) and *G_F_* (*t*), indexed by *t*, the time after the liver stage (i.e., *t* = 0 indicates the time when the initial blood-stage parasites *P*_0_(*a*) are established; mathematically *P* (*a,* 0) = *P*_0_(*a*)). The number of asexual parasites is given as follows:

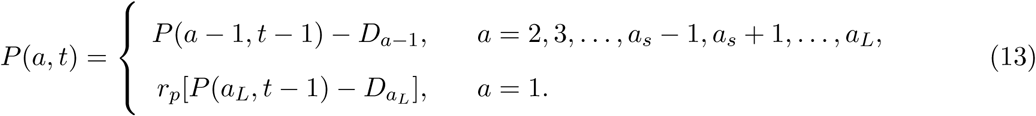

where *a_s_*is the age at which sexual commitment occurs, and *r_p_* is the parasite replication rate. Note that *r_p_* is distinct from the parasite multiplication rate (PMR). The PMR is the ‘net replication rate’, accounting for the increase in parasite numbers due to replication, *r_p_*, and the decrease in parasite numbers due to parasite death or sexual commitment. The PMR is calculated as *r_p_*(1 *− δ_p_*Δ*t*)*^aL/^*^Δ^*^t^*(1 *− f*), where the term (1 *− δ_p_*Δ*t*)*^aL/^*^Δ^*^t^*(1 *− f*) gives the fraction of surviving asexual parasites after death and sexual conversion per replication cycle, *δ_p_* is the rate of natural death of asexual and sexually committed parasites, and *f* is the sexual conversion rate. The number of asexual parasites that die during each time step *D_a_* is sampled from Poi(*P* (*a −* 1*, t −* 1)(*δ_p_* + *k̅_DHA_* + *k̅_P PQ_*)Δ*t*), where *k̅_DHA_* and *k̅_P PQ_* are the average killing rates induced by DHA and PPQ, respectively. The average killing rate, *k̅_d_*, for drug *d ∈ {*DHA, PPQ*}* is computed as the mean of the instantaneous killing rates at times *t −* 1 and *t*, i.e., *k̅_d_* = (*k_d_*(*t −* 1) + *k_d_*(*t*)) */*2, where the drug-specific killing rate at time *t*, *k_d_*(*t*), is given by the standard Hill function *k_d_*(*t*) = *k*_max_*_,d_C_d_*(*t*)*^γd^ /* (*C_d_*(*t*)*^γd^* + *EC*_50_*_,d_^γd^*). The drug concentration, *C_d_*(*t*), is simulated from the PK model, *k*_max_*_,d_* is the maximum killing rate, *EC*_50_*_,d_* is the half-maximal effective concentration, and *γ_d_* is the Hill coefficient. Details of the PK model and PD parameters for DHA are provided in [28], and those for PPQ are provided in [29]. Note that DHA concentrations are simulated using a PK model that accounts for artesunate administration and its rapid conversion to the active metabolite DHA. The effects of DHA and PPQ are assumed to be additive, with no synergistic or antagonistic effects, based on an *in vitro* study indicating no interaction between the two drugs [47]. For parasites that undergo sexual commitment (i.e., parasites of age *a_s_*), the following transitions are applied:

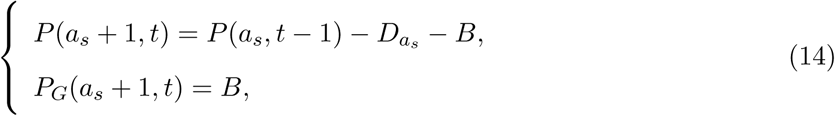

where *B* represents the number of parasites that commit to sexual development, sampled from a binomial distribution, Bin(*P* (*a_s_, t −* 1) *− D_a__s_, f*), and *f* is the probability of sexual commitment for each parasite. The first line of Eq. 14 describes the transition of parasites that continue in the asexual replication cycle, whereas the second line accounts for those that transition into sexually committed forms. Sexually committed parasites continue through the remainder of the replication cycle, modelled by:

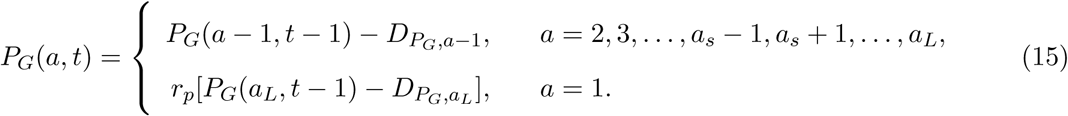

where *D_P__G,a_* represents the number of sexually committed parasites that die during each time step, sampled from Poi(*P_G_*(*a, t −* 1)(*δ_p_* + *k̅_DHA_*)Δ*t*). DHA is assumed to be effective against all parasite stages except mature male and female gametocytes, *G_M_* and *G_F_*, based on *in vitro* experiments [48, 49], whereas PPQ has minimal activity against sexually committed parasites and gametocytes, based on both *in vivo* and *in vitro* experiments [50, 51]. Following sexual commitment, parasites continue their development into immature gametocyte stages, i.e., *G*_1_– *G*_4_, and eventually mature into circulating male and female gametocytes, *G_M_* and *G_F_*. The dynamics of immature gametocytes *G*_1_–*G*_4_ are modelled by:

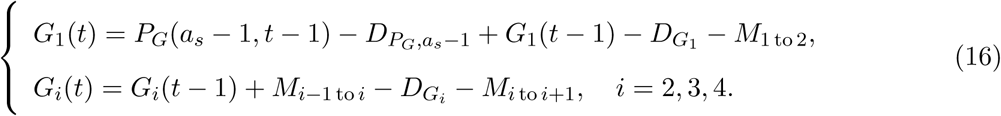

where *D_G__i_* represents the number of stage *i* gametocytes that die during each time step, sampled from Poi(*G_i_*(*t −* 1)(*δ_g_* + *k̅_DHA_*)Δ*t*), and *δ_g_* is the death rate of immature gametocytes. The number of stage-*i* gametocytes maturing into stage-(*i* + 1) gametocytes *M_i_* _to_ *_i_*_+1_ is sampled from Poi(*G_i_*(*t −* 1)*m*Δ*t*) for *i* = 1, 2, 3, 4, where *m* is the rate of gametocyte maturation. Male and female gametocytes develop after stage IV [2] and their dynamics are modelled by:

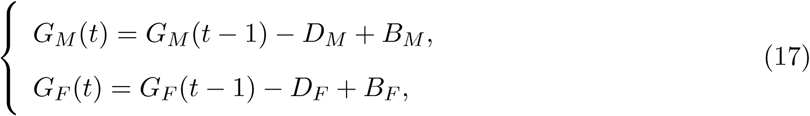

where *D_M_* represents the number of male gametocytes that die during each time step, sampled from Poi(*G_M_* (*t −* 1)*δ_gc_*(1 + *βd_PQ_k̅_PQ_*) Δ*t*), and *δ_gc_* is the natural death rate of circulating gametocytes, *d_PQ_* is the dose of primaquine, *k̅_PQ_* is the average killing rate of primaquine against mature male gametocytes, and *β* is the slope of the linear relationship between dose, *d_PQ_*, and *k̅_PQ_*. Similarly, the number of female gametocytes that die during each time step *D_F_* is sampled from Poi(*G_F_* (*t −* 1)*δ_gc_*(1 + *βd_PQ_k̅_PQ_*)Δ*t*). The average killing rate *k̅_PQ_* is approximated by the average of *k_PQ_*(*t −* 1) and *k_PQ_*(*t*), where *k_PQ_*(*t*) = *E*_max_*C_PQ_*(*t*)*^γ^/*(*C_PQ_*(*t*)*^γ^* + *EC^γ^*). The effective concentration of primaquine *C_PQ_*(*t*) at time *t* is simulated using the primaquine PK model provided in [46], where *E*_max_ is the maximum killing effect of primaquine on circulating gametocytes, *EC*_50_ is the half-maximal effective concentration of primaquine, and *γ* is the Hill coefficient of primaquine. Note that in [46], the PK–PD model of primaquine does not explicitly distinguish between male and female gametocytes. In our model, we directly adopt the PK–PD model and parameter values from [46] and assume that primaquine exerts the same killing effect on both male and female gametocytes. The number of immature gametocytes that become male gametocytes *B_M_*is sampled from a binomial distribution, Bin(*M*_4to5_*, f_G_*), where *f_G_* is the probability that a sequestered *G*_4_ gametocyte develops into a male gametocyte. The number of immature gametocytes that become female gametocytes *B_F_* is given by *M*_4to5_ *− B_M_*.

Ring-stage parasitemia is calculated as 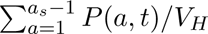, corresponding to the density of circulating parasites (Fig. 1, solid-boundary compartments in the schematic diagram of the within-host model). Parasites sequestered in tissues (Fig. 1, dashed-boundary compartments in the schematic diagram of the within-host model) are not measurable in peripheral blood samples. Total gametocytemia is calculated as (*G_M_* (*t*) + *G_F_* (*t*))*/V_H_*. Infectiousness, denoted by *P_M_* (*t*), is defined as the probability that a mosquito taking a blood meal from an individual at time *t* develops zygotes. This probability is determined by the male and female gametocytes within the individual, *G_M_* (*t*) and *G_F_* (*t*), through the human-to-mosquito transmission model described by Eqs. 4–7. *P_M_* (*t*) is mathematically formulated as follows:

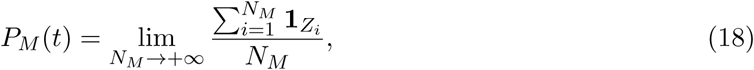

where *N_M_* is the number of mosquitoes that take a blood meal. Due to practical limitations, mosquito feeding experiments typically use tens of mosquitoes to obtain an approximation of infectiousness [46]. However, in model simulations, *N_M_* should take a much larger value to provide a more precise estimate of the probability of transmission when evaluating the effect of primaquine. Therefore, we set *N_M_* =100,000 in all model simulations.

To simulate asymptomatic infections, when ring-stage parasitemia reaches a set-point level of 10^5^ [52, 53], the parasite replication number *r_p_*in Eqs. 13–15 is adjusted to *r_p_*= 1*/*(1 *− δ_p_*Δ*t*)^42^*^/^*^Δ^*^t^/*(1 *− f*), such that the PMR is then equal to 1, which yields cyclical asexual parasitemia around the set-point level, mimicking the involvement of immune responses in controlling parasite growth and producing parasite dynamics observed in endemic areas [54] (see the representative trajectory shown in Fig. 1). The duration of asymptomatic infections (denoted by *T_A_*) typically ranges from several days to several months [55, 56]. We assume that each asymptomatic individual no longer has circulating and sequestered parasites by the end of the period of infection. For symptomatic infections, individuals are assumed to be managed through ongoing routine case management, whereby they seek clinical care and receive treatment when parasitemia reaches approximately 10^9^ parasites/mL [57]. This baseline treatment pathway is included in all simulations, including the MDA scenarios, to represent the continued availability of routine case management. ACT is administered according to WHO guidelines [1], with 120 mg DHA and 960 mg PPQ given at 0, 24, and 48 hours after treatment initiation, i.e., the time at which parasitemia reaches 10^9^. The PK profile is simulated for an individual with a body weight of 60 kg.

### Evaluation of the population-level effects of primaquine

We simulated the population-level effects of primaquine under two intervention settings: mass drug administration (MDA) and routine case management. To simulate the MDA campaign, the agent-based model was initialised by introducing a single infected individual into a malaria-free population and then simulated forward in time until the transmission dynamics reached a steady endemic level. This steady state was then used as the common starting point for comparison, after which an MDA campaign comprising three rounds at monthly intervals, with *c* = 90% population coverage in each round, was simulated, consistent with the design of Deng et al. [19]. Simulations were conducted under two treatment regimens: ACT alone, and ACT combined with a single 0.25 mg/kg dose of primaquine. Participation across MDA rounds was governed by an adherence parameter, *η*. At the start of the campaign, a fixed proportion *cη* of the total population was selected to receive treatment in all three rounds. In each round, an additional proportion *c*(1*−η*) was sampled independently from the remaining population, ensuring that the overall coverage remained *c* across all rounds. We considered *η* = 1, 0.5 and 0, representing full adherence, half adherence, and independent participation across rounds, respectively. For each combination of treatment regimen and participation scenario, 100 stochastic realisations were simulated. To evaluate the effect of primaquine under routine case management, the model was initialised with a single infected individual and simulated under ACT alone case management until it reached a steady endemic level. From this steady state, we simulated two scenarios: continued ACT alone case management and ACT case management with co-administered 0.25 mg/kg primaquine. In total, 100 realisations were performed.

### Model parameters

All model parameters are listed in Table 1. The human parameter values are chosen from [29]. The standard deviation (SD) of the initial parasite age distribution (*σ*) is chosen from [58]. The value for the human blood volume (*V_H_*) is chosen based on [59]. The value for the fraction of sequestered *G*_4_ gametocytes developing into male gametocytes (*f_G_*) is chosen based on [60]. The parameter values for the PK–PD model of primaquine and the death rate of circulating gametocytes are chosen from [46]. The value for the blood meal volume taken by a mosquito (*V*) is chosen based on [61]. The rationale for the value chosen for the fraction of the total human blood volume localised to the site of a mosquito bite (*f_e_*) is provided in [27]. The value for the male gamete-to-gametocyte ratio (*ρ*) and the probability of successful fertilization for each pair of gametes (*p_f_*) are chosen from [27]. The value for the mosquito biting rate *b* is chosen from [45]. The value for the probability of mosquito-to-human transmission *P_MH_* is chosen from [62]. The value for the natural death rate of mosquitoes *µ_v_* is chosen from [63]. The parameter value for the progression rate from exposed to infectious mosquitoes *k_v_* is chosen from [64]. The parameter value for the period of liver-stage infection *l_h_* is chosen from [65]. The parameter value for the proportion of asymptomatic infections *θ* is chosen based on [42,43]. The parameter value for the duration of asymptomatic infection *T_A_*is chosen based on [55]. The human population size is assumed to be *N_H_* = 1000. Based on a human biting rate of approximately 4 bites per person per day in a low-transmission setting [66] and a mosquito biting rate of *b* = 0.4 bites per mosquito per day [45], the implied mosquito-to-human ratio is 10:1, yielding a mosquito population size of *N_V_* =10,000.

**Table 1.** Details of parameters, descriptions, units, values and reference.

| Parameter | Description | Unit | Value | Reference |
| --- | --- | --- | --- | --- |
| $P_{\text{init}}$ | Initial asexual parasite concentration | parasites/mL | 0.08 | [29] |
| $V_H$ | Human blood volume | mL | 4000 | [59] |
| $\mu$ | Mean of the initial age distribution | h | 7 | [29] |
| $\sigma$ | SD of the initial age distribution | h | 8 | [58] |
| $r_p$ | Replication number | unitless | 71 | [29] |
| $f$ | Fraction of parasites entering sexual development | unitless | 0.006 | [29] |
| $\delta_p$ | Death rate of asexual and sexual parasites | $\text{h}^{-1}$ | 0.03 | [29] |
| $\delta_g$ | Death rate of sequestered gametocytes | $\text{h}^{-1}$ | 0.0004 | [29] |
| $\delta_{gc}$ | Death rate of circulating gametocytes | $\text{h}^{-1}$ | 0.002 | [46] |
| $m$ | Maturation rate of gametocytes | $\text{h}^{-1}$ | 0.02 | [29] |
| $f_g$ | Fraction of sequestered gametocytes becoming male gametocytes | unitless | 0.2 | [51] |
| $d_{PQ}$ | Dose of primaquine | mg/kg | 0.25 | [46] |
| $E_{\text{max}}$ | Maximum killing effect of primaquine on gametocytes | $\text{h}^{-1}$ | 84 | [46] |
| $EC_{50}^{574}$ | Half-maximum effective primaquine concentration | nmol/mL | 0.0001 | [46] |
| $\gamma$ | Hill coefficient of primaquine | unitless | 1 | [46] |
| $\beta$ | Fold change of primaquine | per 1 mg/kg | 2.44 | [46] |
| $V$ | Blood meal volume taken by mosquito | $\mu\text{l}$ | 3.4 | [61] |
| $f_e$ | Fraction of total human blood volume localized to the site of a mosquito bite | unitless | 0.00025 | [27] |
| $\rho$ | Male gamete-to-gametocyte ratio | unitless | 0.8 | [27] |
| $p_f$ | Probability of successful fertilization | unitless | 0.03 | [27] |
| $b$ | Biting rate per mosquito | $\text{Day}^{-1}$ | 0.4 | [45] |
| $P_{MH}$ | Probability of mosquito-to-human transmission | unitless | 0.55 | [62] |
| $\mu_v$ | Death rate of mosquitoes | $\text{Day}^{-1}$ | 1/12 | [63] |
| $k_v$ | Mosquito latent rate | $\text{Day}^{-1}$ | 1/12 | [64] |
| $l_h$ | Latent time for liver stage infection | Day | 7 | [67] |
| $\theta$ | Proportion of asymptomatic infections | unitless | 0.05 | [42, 43] |
| $T_A$ | Duration of asymptomatic infections | Day | 194 | [55] |

## Data Availability

No empirical data were collected or analysed in this study. All the model parameters were obtained from previously published studies.

## Code availability

All the code used for model analysis and result visualization is publicly available at https://github.com/XSSUN2/Agent-based-transmission-model. All analyses were performed in R (version 4.2.1).

## Acknowledgments

We thank the Australian Research Council (ARC) for supporting this research through a Discovery Project (DP210101920) and a Laureate Fellowship awarded to JMM (FL240100126) which supports XS, PC and JMM. JAS was supported by a National Health and Medical Research Council (NHMRC) Investigator Grant (2042554). Computer simulations were conducted on the Nectar Research Cloud, a collaborative Australian research platform supported by the National Collaborative Research Infrastructure Strategy (NCRIS).

## Author contributions

XS, JAS, JMM and PC contributed to conceptualization. XS contributed to formal analysis. XS, JMM and PC contributed to methodology. JMM and PC contributed to supervision. XS contributed to visualization. XS, JMM and PC contributed to writing of the original draft manuscript. XS, JAS, JMM and PC contributed to review and editing of the final manuscript.

## Supplementary Materials

**Supplementary Figure 1.**
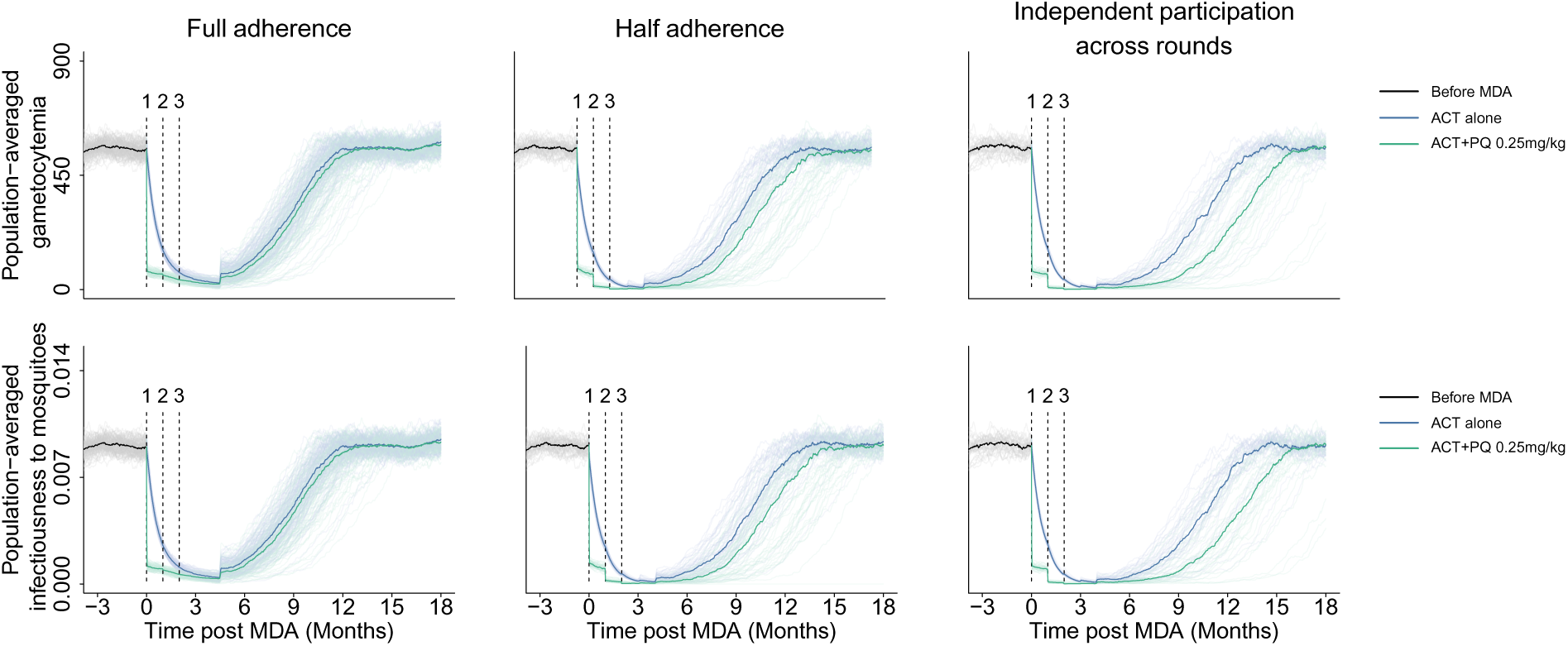
Effect of adding a single dose of 0.25 mg/kg primaquine to ACT on population-averaged gametocytemia and infectiousness to mosquitoes during MDA under different adherence scenarios.

## References

[1] World Health Organization. WHO guidelines for malaria, 13 August 2025 (Geneva: World Health Organization, 2025). URL 10.2471/B09514. Licence: CC BY-NC-SA 3.0 IGO.

[2] Josling, G. A. & Llińas, M. Sexual development in Plasmodium parasites: knowing when it’s time to commit. Nature Reviews Microbiology 13, 573–587 (2015).

[3] Okello, G. & Aucamp, M. Malaria: a review on its current epidemiological status and management strategies. Malaria Journal 25, 132 (2026).

[4] Birkholtz, L.-M., Alano, P. & Leroy, D. Transmission-blocking drugs for malaria elimination. Trends in Parasitology 38, 390–403 (2022).

[5] Dicko, A. et al. Primaquine to reduce transmission of *Plasmodium falciparum* malaria in Mali: a single-blind, dose-ranging, adaptive randomised phase 2 trial. The Lancet. Infectious diseases 16, 674–684 (2016).

[6] Dicko, A. et al. Efficacy and safety of primaquine and methylene blue for prevention of *Plasmodium falciparum* transmission in Mali: a phase 2, single-blind, randomised controlled trial. The Lancet. Infectious Diseases 18, 627–639 (2018).

[7] Stone, W. et al. Single low-dose tafenoquine combined with dihydroartemisinin–piperaquine to reduce *Plasmodium falciparum* transmission in Oue-lessebougou, Mali: a phase 2, single-blind, randomised clinical trial. The Lancet Microbe 3, e336–e347 (2022).

[8] Yilma, D. et al. Safety and efficacy of single-dose primaquine to interrupt *Plasmodium falciparum* malaria transmission in children compared with adults: a systematic review and individual patient data meta-analysis. The Lancet Infectious Diseases 25, 965–976 (2025).

[9] Eziefula, A. C. et al. Single dose primaquine for clearance of *Plasmodium falciparum* gametocytes in children with uncomplicated malaria in Uganda: a randomised, controlled, double-blind, dose-ranging trial. The Lancet Infectious Diseases 14, 130–139 (2014).

[10] Goņcalves, B. P., et al. Single low dose primaquine to reduce gametocyte carriage and *Plasmodium falciparum* transmission after artemether-lumefantrine in children with asymptomatic infection: a randomised, double-blind, placebo-controlled trial. BMC Medicine 14, 70 (2016).

[11] Stone, W. et al. Pyronaridine–artesunate or dihydroartemisinin–piperaquine combined with single low-dose primaquine to prevent *Plasmodium falciparum* malaria transmission in oúelesśebougou, Mali: a four-arm, single-blind, phase 2/3, randomised trial. The Lancet. Microbe 3, e41–e51 (2021).

[12] Mwaiswelo, R. O. et al. A single low dose of primaquine is safe and sufficient to reduce transmission of *Plasmodium falciparum* gametocytes regardless of cytochrome P450 2d6 enzyme activity in Bagamoyo district, Tanzania. Malaria Journal 21, 84 (2022).

[13] Stepniewska, K. et al. Safety of single-dose primaquine as a *Plasmodium falciparum* gametocytocide: a systematic review and meta-analysis of individual patient data. BMC Medicine 20, 350 (2022).

[14] Graves, P. M., Gelband, H. & Garner, P. Primaquine or other 8-aminoquinoline for reducing P. falciparum transmission. Cochrane Database of Systematic Reviews 6, CD008152 (2014).

[15] Ye, T. et al. Single low-dose primaquine for malaria control in Africa: a systematic review of safety, efficacy and implementation barriers. BMJ Global Health 10, e020264 (2025).

[16] Aung, P. L. et al. Factors hindering coverage of targeted mass treatment with primaquine in a malarious township of northern Myanmar in 2019–2020. Scientific Reports 13, 5963 (2023).

[17] Morris, U. et al. A cluster randomised controlled trial of two rounds of mass drug administration in Zanzibar, a malaria pre-elimination setting—high coverage and safety, but no significant impact on transmission. BMC Medicine 16, 215 (2018).

[18] Kagaya, W. et al. Malaria resurgence after significant reduction by mass drug administration on Ngodhe island, Kenya. Scientific Reports 9, 19060 (2019).

[19] Deng, C. et al. Large-scale artemisinin–piperaquine mass drug administration with or without primaquine dramatically reduces malaria in a highly endemic region of Africa. Clinical Infectious Diseases 67, 1670–1676 (2018).

[20] Maude, R. J. et al. Optimising strategies for *Plasmodium falciparum* malaria elimination in Cambodia: Primaquine, mass drug administration and artemisinin resistance. PLoS ONE 7, e37166 (2012).

[21] Gerardin, J., Eckhoff, P. A. & Wenger, E. A. Mass campaigns with antimalarial drugs: a modelling comparison of artemether-lumefantrine and DHA-piperaquine with and without primaquine as tools for malaria control and elimination. BMC Infectious Diseases 15, 144 (2015).

[22] Lawley, S. D., Gibson, T. B. & Jiao, Z. Editorial: Mathematical modeling of medication nonadherence. Frontiers in Pharmacology 16, 1741041 (2025).

[23] Adhikari, B. et al. Factors associated with population coverage of targeted malaria elimination (TME) in southern Savannakhet province, Lao PDR. Malaria Journal 16, 424 (2017).

[24] von Seidlein, L. et al. The impact of targeted malaria elimination with mass drug administrations on falciparum malaria in Southeast Asia: A cluster randomised trial. PLoS Medicine 16, e1002745 (2019).

[25] Dyson, L., Stolk, W. A., Farrell, S. H. & Hollingsworth, T. D. Measuring and modelling the effects of systematic non-adherence to mass drug administration. Epidemics 18, 56–66 (2017).

[26] Hardwick, R. J. et al. Individual adherence to mass drug administration in neglected tropical disease control: A probability model conditional on past behaviour. PLoS Neglected Tropical Diseases 15, e0009112 (2021).

[27] Sun, X., Dixon, M. W. A., McCarthy, J. S., McCaw, J. M. & Cao, P. Coupling models of within-human, human-to-mosquito, and within-mosquito malaria parasite dynamics to identify key drivers of malaria transmission. medRxiv (2025). URL https://www.medrxiv.org/content/10.1101/2025.10.28.25339021v1. Preprint.

[28] Das, J. L. et al. Population pharmacokinetic and pharmacodynamic modeling of artemisinin resistance in Southeast Asia. The AAPS Journal 19, 1842–1854 (2017).

[29] Cao, P. et al. Modeling the dynamics of *Plasmodium falciparum* gametocytes in humans during malaria infection. eLife 8, e49058 (2019).

[30] Okombo, J. & Fidock, D. A. Towards next-generation treatment options to combat *Plasmodium falciparum* malaria. Nature Reviews Microbiology 23, 178–191 (2024).

[31] Churcher, T. S. et al. Predicting mosquito infection from Plasmodium falciparum gametocyte density and estimating the reservoir of infection. eLife 2, e00626 (2013).

[32] Dierickx, S. et al. Factors associated with non-participation and non-adherence in directly observed mass drug administration for malaria in the Gambia. PLoS ONE 11, e0148627 (2016).

[33] Fehr, A. E. et al. From informed consent to adherence: factors influencing involvement in mass drug administration with ivermectin for malaria elimination in the Gambia. Malaria Journal 20, 198 (2021).

[34] Rogers, K. J., Vijay, R. & Butler, N. S. Anti-malarial humoral immunity: the long and short of it. Microbes and Infection 23, 104807 (2021).

[35] Ataide, R. et al. Host immunity to Plasmodium falciparum and the assessment of emerging artemisinin resistance in a multinational cohort. Proceedings of the National Academy of Sciences 114, 3515–3520 (2017).

[36] Stuckey, E. M. et al. Modeling the cost effectiveness of malaria control interventions in the highlands of western Kenya. PLoS ONE 9, e107700 (2014).

[37] Okell, L. C. et al. Contrasting benefits of different artemisinin combination therapies as first-line malaria treatments using model-based cost-effectiveness analysis. Nature Communications 5, 5606 (2014).

[38] Crowell, V. et al. Modelling the cost-effectiveness of mass screening and treatment for reducing *Plasmodium falciparum* malaria burden. Malaria Journal 12, 4 (2013).

[39] Eziefula, A. C. et al. Rationale for short course primaquine in Africa to interrupt malaria transmission. Malaria Journal 11, 360 (2012).

[40] Okell, L. C. et al. The potential contribution of mass treatment to the control of *Plasmodium falciparum* malaria. PLoS ONE 6, e20179 (2011).

[41] Griffin, J. T. et al. Reducing *Plasmodium falciparum* malaria transmission in africa: A model-based evaluation of intervention strategies. PLoS Medicine 7, e1000324 (2010).

[42] Tamiru, A., Tolossa, T., Regasa, B. & Mosisa, G. Prevalence of asymptomatic malaria and associated factors in Ethiopia: Systematic review and meta-analysis. SAGE Open Medicine 10, 20503121221088085 (2022).

[43] Kotepui, M., Kotepui, K. U., Masangkay, F. R., Mahittikorn, A. & Wilairatana, P. Prevalence and proportion estimate of asymptomatic Plasmodium infection in Asia: a systematic review and meta-analysis. Scientific Reports 13, 10379 (2023).

[44] Simoy, M. I. & Aparicio, J. P. Ross-macdonald models: Which one should we use? Acta tropica 207, 105452 (2020).

[45] Santos, R. L. C. d., Forattini, O. P. & Burattini, M. N. Laboratory and field observations on duration of gonotrophic cycle of *Anopheles albitarsis* s.l. (diptera: Culicidae) in southeastern Brazil. Journal of Medical Entomology 39, 926–930 (2002).

[46] Chotsiri, P. et al. Mechanistic modeling of primaquine pharmacokinetics, gametocytocidal activity, and mosquito infectivity. Clinical Pharmacology and Therapeutics 111, 676–685 (2021).

[47] Davis, T. M. E. et al. In vitro interactions between piperaquine, dihydroartemisinin, and other conventional and novel antimalarial drugs. Antimicrobial Agents and Chemotherapy 50, 2883–2885 (2006).

[48] Adjalley, S. H. et al. Quantitative assessment of *Plasmodium falciparum* sexual development reveals potent transmission-blocking activity by methylene blue. Proceedings of the National Academy of Sciences 108, E1214–E1223 (2011).

[49] Lelìevre, J., et al. Activity of clinically relevant antimalarial drugs on *Plasmodium fal-ciparum* mature gametocytes in an ATP bioluminescence “transmission blocking” assay. PLoS ONE 7, e35019 (2012).

[50] Bolscher, J. M. et al. A combination of new screening assays for prioritization of transmission-blocking antimalarials reveals distinct dynamics of marketed and experimental drugs. The Journal of antimicrobial chemotherapy 70, 1357–1366 (2015).

[51] Collins, K. A. et al. A controlled human malaria infection model enabling evaluation of transmission-blocking interventions. The Journal of Clinical Investigation 128, 1551–1562 (2018).

[52] Slater, H. C. et al. The temporal dynamics and infectiousness of subpatent *Plasmodium falciparum* infections in relation to parasite density. Nature Communications 10, 1433 (2019).

[53] Goņcalves, B. P., et al. Examining the human infectious reservoir for *Plasmodium falciparum* malaria in areas of differing transmission intensity. Nature Communications 8, 1133 (2017).

[54] Hergott, D. E. B. et al. Assessing the daily natural history of asymptomatic Plasmodium infections in adults and older children in Katakwi, Uganda: a longitudinal cohort study. The Lancet Microbe 5, e72–e80 (2024).

[55] Felger, I. et al. The dynamics of natural Plasmodium falciparum infections. PLOS ONE 7, e45542 (2012).

[56] Roe, M. S., O’Flaherty, K. & Fowkes, F. J. I. Can malaria parasites be spontaneously cleared? Trends in Parasitology 38, 356–364 (2022).

[57] Abdulla, S. M. et al. Baseline data of parasite clearance in patients with falciparum malaria treated with an artemisinin derivative: an individual patient data meta-analysis. Malaria Journal 14, 359 (2015).

[58] Bozdech, Z. et al. The transcriptome of the intraerythrocytic developmental cycle of *Plasmodium falciparum*. PLoS Biology 1, e5 (2003).

[59] Hale, A. J., Ricotta, D. N., Herzig, S. J., William, J. H. & Freed, J. A. A quantitative approach to dilutional anemia. Journal of Hematology 8, 86–87 (2019).

[60] Tadesse, F. G. et al. Gametocyte sex ratio: The key to understanding *Plasmodium falciparum* transmission? Trends in Parasitology 35, 226–238 (2019).

[61] Graumans, W. et al. A mosquito feeding assay to examine Plasmodium transmission to mosquitoes using small blood volumes in 3d printed nano-feeders. Parasites Vectors 13, 401 (2020).

[62] Smith, D. L., Drakeley, C. J., Chiyaka, C. & Hay, S. I. A quantitative analysis of transmission efficiency versus intensity for malaria. Nature Communications 1, 108 (2010).

[63] Matthews, J., Bethel, A. C. & Osei, G. An overview of malarial Anopheles mosquito survival estimates in relation to methodology. Parasites & Vectors 13, 233 (2020).

[64] Stopard, I. J., Churcher, T. S. & Lambert, B. Estimating the extrinsic incubation period of malaria using a mechanistic model of sporogony. PLoS Computational Biology 17, e1008658 (2021).

[65] Soulard, V., et al. *Plasmodium falciparum* full life cycle and Plasmodium ovale liver stages in humanized mice. Nature Communications 6, 7690 (2015).

[66] Doucoure, S. et al. Anopheles arabiensis and Anopheles funestus biting patterns in Dielmo, an area of low level exposure to malaria vectors. Malaria Journal 19, 230 (2020).

[67] Hermsen, C. C. et al. Detection of *Plasmodium falciparum* malaria parasites in vivo by real-time quantitative PCR. Molecular and Biochemical Parasitology 118, 247–251 (2001).

